# Baseline Structural and Functional Magnetic Resonance Imaging Predicts Early Treatment Response in Schizophrenia with Radiomics Strategy

**DOI:** 10.1101/2020.02.06.20020784

**Authors:** Long-Biao Cui, Yu-Fei Fu, Lin Liu, Yongbin Wei, Xu-Sha Wu, Yi-Bin Xi, Hua-Ning Wang, Wei Qin, Hong Yin

**Affiliations:** Department of Clinical Psychology, School of Medical Psychology, Fourth Military Medical University, Xi’an, China; Department of Psychiatry, Xijing Hospital, Fourth Military Medical University, Xi’an, China; Department of Radiology, Xijing Hospital, Fourth Military Medical University, Xi’an, China; School of Life Sciences and Technology, Xidian University, Xi’an, China; Sixth Hospital/Institute of Mental Health and Key Laboratory of Mental Health, Peking University, Beijing, China; Connectome Lab, Department of Complex Trait Genetics, Center for Neurogenomics and Cognitive Research, Amsterdam Neuroscience, Vrije Universiteit Amsterdam, Amsterdam, The Netherlands

**Keywords:** schizophrenia, antipsychotics, early treatment response, prediction, magnetic resonance imaging, radiomics

## Abstract

Multimodal neuroimaging features might enable accurate classification and provide personalized treatment options in psychiatric domain. We conducted a retrospective study to investigate whether structural and functional features for predicting response to overall treatment of schizophrenia at the end of the first or a single hospitalization and in addition cross validate the results. This structural and functional magnetic resonance imaging (MRI) study included 85 and 63 patients with schizophrenia at baseline in dataset 1 and 2, respectively. After treatment, patients were classified as responders and non-responders. Features of gray matter and functional connectivity were extracted. Radiomics analysis was used to explore the predictive performance. Prediction models were based on structural features, functional features, and combined features. We found that the prediction accuracy was 80.38% (sensitivity: 87.28%; specificity 82.47%) for the model using functional features, and 69.68% (sensitivity: 83.96%; specificity: 72.41%) for the model using structural features. Our model combined both structural and functional features accurately predicted 92.04% responder and 80.23% non-responders to overall treatment, with an accuracy of 85.03%. These results highlight the power of structural and functional MRI-derived radiomics features to predict early response to treatment in schizophrenia. Prediction models of the very early treatment response in schizophrenia could augment effective therapeutic strategies.

## Introduction

Schizophrenia is a severe mental disorder that ranks among the major conditions contributing the global burden of disease and is regarded as a global public-health challenge. Currently, an obvious clinical obstacle for schizophrenia is a lack of personalized treatment. Driven by the need for better management of schizophrenia, as well as advances in neuroimaging,^1^ a quest for accurate prediction of early treatment response^2, 3^ was noted in psychiatric domain.

Longer duration of untreated psychosis is associated with poorer outcome,^4^ including social functioning, response to treatment, and physical illnesses, making a compelling argument for treating patients with schizophrenia or other psychotic disorders as soon as possible.^5^ On the contrary, shortening the interval between the first onset of schizophrenia and the start of intervention leads to better outcomes.^6^ Early intervention service can save young lives, and young people may benefit from disease-modifying strategies applied early.^7^ The notion of emphasizing the early intervention should also be paralleled by predicting response at early stage, because predictive markers can be helpful in treatment selection. Emerging neuroimaging studies indicate that structural and functional magnetic resonance imaging (MRI) techniques may be able to predict the response of treatments in schizophrenia, as well as first-episode or acute schizophrenia (see Cui et al for review).^3^

To this end, radiomics is gaining significance in psychiatric research, enabling imaging data to improve predictive accuracy within clinical decision support.^8^ Radiomics is a powerful tool in psychiatry and is evolving rapidly, producing robust for schizophrenia^9^ and attention deficit hyperactivity disorder.^10^ In processing, the features after rigorous preprocessing of each imaging modal were selected as radiomics features. Here, prediction model was based on all of the features of training, without any subjective selection. Each feature after dimensionality reduction contributes to the prediction of clinical efficacy. In contrast, conventional prediction methods usually calculate the inter-group differences of imaging data and then predict some clinical outcome through correlation analysis. Due to individual differences of schizophrenia, after comparison with correction between different groups, some characteristics may not be detected, which may serve the classification and prediction. Promisingly, a valid approach by means of functional connectivity to diagnose schizophrenia has been developed using radiomics strategy with an accuracy of 87%.^9^ Its increasing importance in medical imaging creates an ideal situation for application of radiomics in neuroradiology where there is no “lesion” but there are “features” for mental disorders, i.e., brain structure and connectome derived from MRI.^11, 12^ Future research needs to integrate and optimize them to achieve an accurate prediction for individualized clinical management of schizophrenia.

Given such a background, we aimed to investigate the psycho-radiomic application using structural morphology and functional connectivity to predict the response to overall treatment of schizophrenia at the end of the first or a single hospitalization. The mean length of stay in hospital is around 2-3 weeks (arranging from 17.2 days to 20.3 days) for patients in the current study. This period is of clinical importance because patients will usually demonstrate whether they respond to an initial antipsychotic medication or not in the first 2 weeks (see the Appendix for review), also, response of the first 2-4 weeks of treatment is remarkably predictive of long-term response.^13, 14^ In addition, we cross-validated our results via two independent cohorts. We hypothesized that this strategy can be harnessed via structural and functional MRI at baseline and leveraged through radiomics approach to aid prediction of early response to anti-psychotic treatment. Imaging-based model to predict early response contributes to the precise medicine in psychiatric domain.

## Methods

### Participants

Two independent datasets were included in this study, and a partial sample in this study have been investigated in Cui et al,^9, 15^ which report the inclusion and exclusion criteria in details. The dataset 1 included 85 patients with schizophrenia, which were collected between May 2011 through September 2013. The structural clinical interview for Diagnostic and Statistical Manual of Mental Disorders, Fourth Edition, Text Revision (DSM-IV-TR) were used, and consensus diagnoses were made using all the available information. Each patient was assessed by using the Positive and Negative Syndrome Scale (PANSS) at the time of imaging. The dataset 2 included 63 patients with schizophrenia spectrum disorder, including schizophrenia (n = 34), schizophreniform disorder (n = 22), and brief psychotic disorder (n = 7). Patients were diagnosed according to DSM, Fifth Edition (DSM-5) between April 2015 through December 2017, with no more than two weeks of cumulative exposure to antipsychotics. Those with illness duration of fewer than six months were assessed through follow-up clinical evaluations. Patients who were diagnosed with schizophrenia during follow-up were included in the current study. This study was approved by the local ethics committee of Xijing Hospital. All participants (or their parents for those under age of 18 years) gave written informed consent after a full description of the aims and design of the study.

In this study, the first hospitalization refers to first episode patients, and a single hospitalization refers to non-first episode patients. The majority of patients received second-generation antipsychotics, including risperidone (61.4% in dataset 1; 57.1% in dataset 2), olanzapine (27.3%; 28.6%), ziprasidone (8.0%; 1.6%), quetiapine (12.5%; 1.6%), paliperidone (12.5%; 17.5%), aripiprazole (4.5%; 9.5%), amisulpride (2.3%; 6.3%), and clozapine (0.0%; 1.6%). The minority of patients received first-generation antipsychotics, including haloperidol (19.3%; 15.9%), chlorpromazine (3.4%; 1.6%), perphenazine (2.3%; 0.0%), and sulpiride (1.1%; 1.6%). A total of 18 patients in dataset 1 and 21 patients in dataset 2 were treated with electroconvulsive therapy (ECT). Moreover, 58 patients in dataset 2 underwent repetitive transcranial magnetic stimulation (rTMS). Treatment response before discharging was assessed using percentage change of symptoms based on PANSS. Responders were defined as 30% reduction in PANSS total scores traditionally used.^16, 17^ Table 1 provides further details on the two patient populations.

**Table 1.** Sample Demographics

| Characteristic | Dataset 1 |  |  | Dataset 2 |  |  |
| --- | --- | --- | --- | --- | --- | --- |
|  | Respond | Non-respond | *P* | Respond | Non-respond | *P* |
|  | ers | ers | valu | ers | ers | valu |
|  | (n = 47) | (n = 38) | es | (n = 41) | (n = 22) | es |
| Age (y) | 25.1 ± | 26.0 ± 7.0 | 0.87 | 21.9 ± | 26.0 ± 9.1 | 0.02 |
|  | 5.7 |  | 7 | 5.5 |  | 8 |
| Gender (M/F) | 26/21 | 22/16 | 0.81 | 27/14 | 11/11 | 0.22 |
|  |  |  | 2 |  |  | 0 |
| Education | 13.2 ± | 12.8 ± 1.8 | 0.35 | 12.1 ± | 12.5 ± 3.4 | 0.60 |
| level (y) | 1.8 |  | 4 | 2.7 |  | 1 |
| Duration of | 18.2 ± | 27.5 ± 33.6 | 0.15 | 11.9 ± | 17.1 ± 28.3 | 0.42 |
| illness (mon) | 23.0 |  | 8 | 14.7 |  | 8 |
| FE/NFE | 30/19 | 21/17 |  | 36/5 | 19/3 |  |
| PANSS score |  |  |  |  |  |  |
| at baseline |  |  |  |  |  |  |
| Total score | 97.6 ± | 91.1 ± 14.3 | 0.08 | 83.0 ± | 87.9 ± 12.2 | 0.33 |
|  | 20.0 |  | 8 | 21.8 |  | 3 |
| Positive score | 24.3 ± | 23.1 ± 7.9 | 0.45 | 22.2 ± | 21.3 ± 5.5 | 0.57 |
|  | 6.4 |  | 1 | 6.2 |  | 3 |
| Negative | 23.2 ± | 23.19 ± 6.7 | 0.97 | 19.2 ± | 22.2 ± 8.7 | 0.13 |
| score | 9.4 |  | 6 | 6.7 |  | 3 |
| General | 50.1 ± | 44.9 ± 7.7 | 0.00 | 43.7 ± | 44.4 ± 6.4 | 0.72 |
| psychopathol | 9.6 |  | 8 | 9.5 |  | 0 |
| ogy score |  |  |  |  |  |  |
| PANSS score |  |  |  |  |  |  |
| at discharging |  |  |  |  |  |  |
| Total score | 66.83 ± | 81.0 ± 11.6 | < | 54.0 ± | 79.3 ± 10.9 | < |
|  | 15.5 |  | 0.00 | 11.2 |  | 0.00 |
|  |  |  | 1 |  |  | 1 |
| Positive score | 15.8 ± 4.5 | 19.8 ± 5.8 | 0.00 | 12.7 ± 4.4 | 18.3 ± 4.4 | < 0.00 |
|  |  |  | 1 |  |  | 1 |
| Negative score | 16.2 ± 6.0 | 21.0 ± 6.0 | < 0.00 | 11.7 ± 3.6 | 21.3 ± 6.7 | < 0.00 |
|  |  |  | 1 |  |  | 1 |
| General psychopathology score | 34.8 ± 8.2 | 40.2 ± 6.0 | 0.00 | 29.5 ± 6.6 | 39.7 ± 4.2 | < 0.00 |
|  |  |  | 1 |  |  | 1 |
| Stay in hospital (d) | 20.3 ± 11.0 | 17.2 ± 7.8 | 0.15 | 20.0 ± 8.7 | 17.4 ± 7.1 | 0.24 |
| Treatment without/with ECT | 35/12 | 32/6 | 0.27 | 26/15 | 16/6 | 0.45 |
|  |  |  | 4 |  |  | 5 |
| Antipsychotic dose, mg/d<sup>a</sup> | 11.4 ± 4.7 | 10.9 ± 4.9 | 0.62 | 12.0 ± 5.5 | 11.5 ± 6.5 | 0.73 |
|  |  |  | 5 |  |  | 5 |
| Changes in PANSS score, % | 47 ± 14 | 16 ± 11 | < 0.00 | 57 ± 17 | 14 ± 18 | < 0.00 |
|  |  |  | 1 |  |  | 1 |

ECT, electroconvulsive therapy; FE, first episode; NFE, non-first episode; PANSS, Positive and Negative Syndrome Scale.
<sup>a</sup>Dose of current antipsychotic medication was converted to Defined Daily Dose (DDD) (WHO Collaborating Centre for Drug Statistics Methodology, 2014).

**Table 2.**
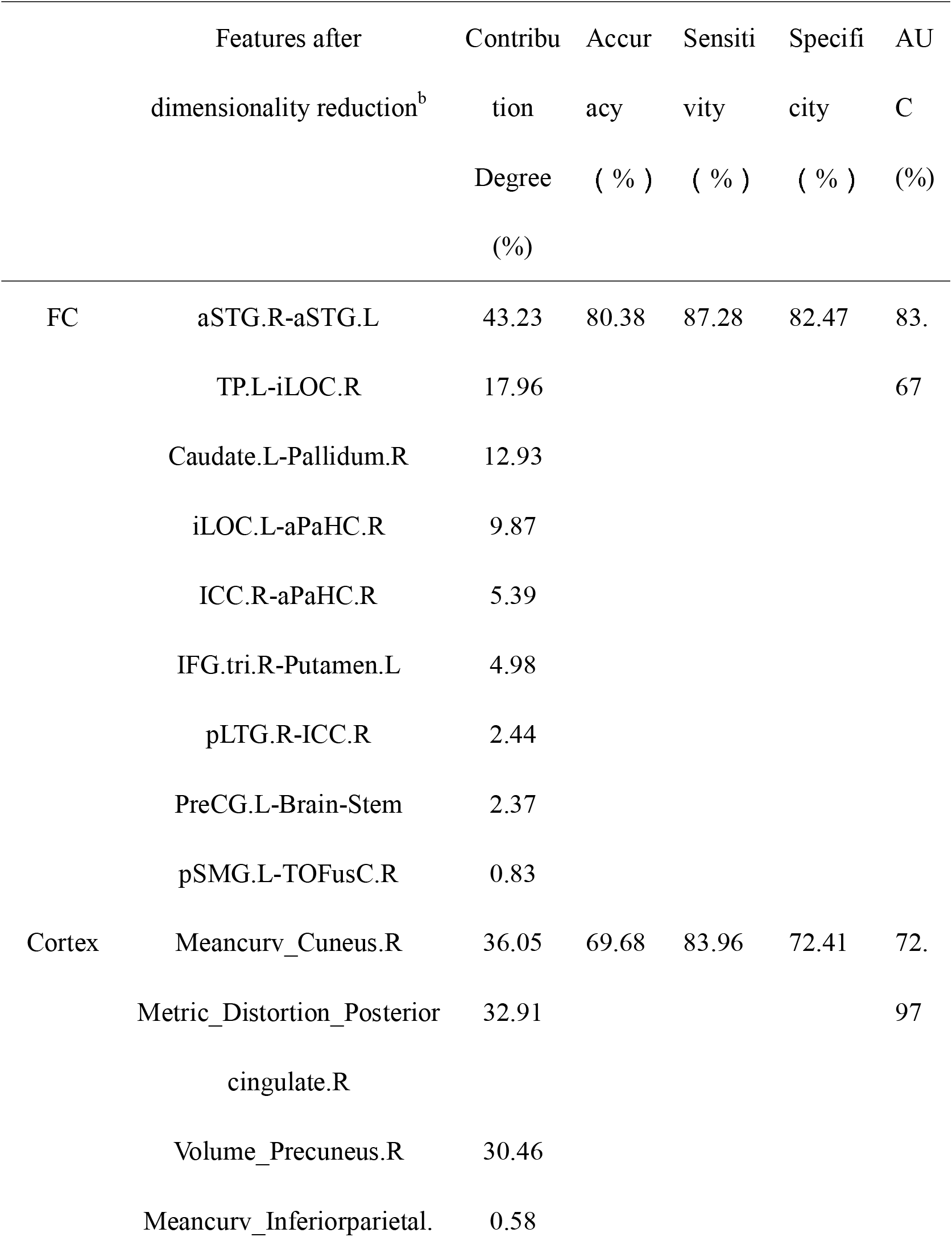
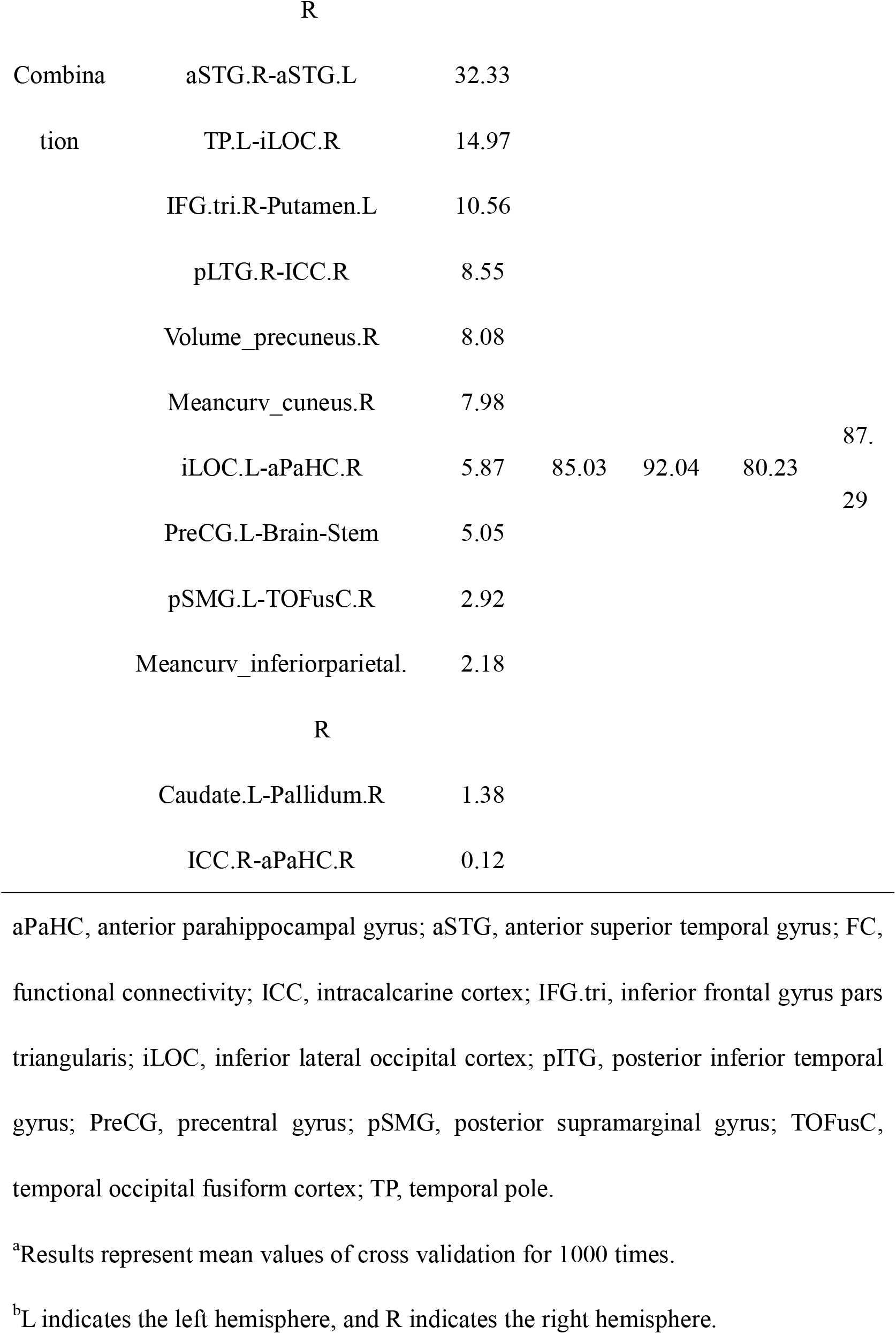
Results of Cross Validation^a^

A part of overlapping participants was used elsewhere: 50 of the 85 patients in dataset 1 and 36 of the 63 patients in dataset 2 have been previously reported.^3, 9, 12, 18^ The previous studies dealt with neural substrates of auditory verbal hallucinations,^18^ disease definition ^9^ and prediction of response to treatment,^3^ and connectome ^12^ using functional or structural imaging whereas in this manuscript we report on the predictive capacity of functional and structural MRI after early treatment for schizophrenia via radiomics.

### Image Acquisition

High-resolution structural and functional imaging was acquired on a Siemens 3.0 T scanner for dataset 1 and a GE 3.0 T scanner for dataset 2 using protocols published previously ^9^. Dataset 1 and 2 were combined for the following analysis.

### Data Preprocessing

Functional images of two datasets were preprocessed using the CONN toolbox (functional connectivity toolbox v17.f, https://web.conn-toolbox.org/). Firstly, the first 10 volumes were excluded to avoid the interference of magnetic field and ensure the signal to reach equilibrium. To remove spurious sources of variance, linear detrending was performed for time series of all of the regions of interest (ROI). Subjects were excluded if any translation or rotation parameters of head movements were exceeded ± 1 mm and/or ± 1°. The corrected functional images were firstly co-registered to each subject’s T1 images without re-slicing. Then, T1 images were normalized to the Montreal Neurological Institute (MNI) space using the Statistical Parametric Mapping (SPM) software, which generated a transformed matrix from native space to the MNI space. The resulted transformed matrix was used to normalize functional images to the MNI space. Next, the scrubbing procedure implemented in the Artifact Detection Tools (ART) (http://www.nitrc.org/projects/artifact_detect.html) was applied to minimize image artifacts due to head movement. Six head motion parameters and their first-level derivative, the averaged signal of cerebrospinal fluid and white matter, and the scrubbing signal from the time series were regressed out. Band-pass filtering (0.01–0.1 Hz) and smoothing with a 6 mm full width at half maximum Gaussian kernel were performed.

T1 Sequence image processing was performed using the Freesurfer image analysis suite (version 6.0, http://surfer.nmr.mgh.harvard.edu/). Briefly, preprocessing was performed with the following steps: (a) skull stripping, (b) normalization to a standard anatomical template, (c) correction for bias-field homogeneity, (d) segmentation of subcortical white matter and deep gray matter volumetric structures, (e) gray-white mater boundary tessellation and a series of deformation procedures which consist of surface inflation, (f) registration to a spherical atlas and parcellation of the cerebral cortex into units based on the gyral and sulcal structures.

### Features Construction

#### Functional connectivity features construction

A complete brain parcellation including 91 cortical areas and 15 subcortical areas from the FSL Harvard-Oxford Atlas was used to segment the whole brain into 106 non-cerebellar anatomical ROI. For each subject, each ROI’s time series was extracted as the average time series across all voxels within that region. Finally, Pearson correlation coefficients were calculated between each pair of preprocessed ROI time series, and a temporal correlation matrix of the size of 106 × 106 was obtained for each subject.

#### Cortical features construction

Due to cortical abnormalities implicated in schizophrenia, cortical neuroanatomical features were used in the current study. The information which collected from preprocessing was used for calculating 408 structural measures morphological features, including volumetric (68 measures of cortical thickness, surface area, and gray matter regional volume) and geometric (68 measures of mean curvature, metric distortion, and sulcal depth) based on Desikan-Killiany Atlas.

### Features Selection

We used a 10-folds cross validation-based Least Absolute Shrinkage and Selection Operator (CV-LASSO) method to further select features (see the Appendix for review). Briefly, 148 subjects were randomly separated into 10 groups. Each time, one group in turn was excluded from the dataset, and the LASSO method with mean of square error (MSE) as the cost function was used on the remaining nine groups to narrow down the initial 5565 functional connectivity features and 408 cortical morphology features into the most important features according to the MSE + 1 SE criteria. This step was repeated 10 times, which resulted in 10 different groups of selected features. Finally, the functional connections that were included in the selected feature group at each step (i.e., 10 groups) were selected as LASSO features for further analysis.^9^

### Classification Model

In our study, the differences of functional connectivity and cortical features between patients in the two datasets were calculated, and no significant difference was found before we selected the prediction model (supplementary table 1 in Appendix). We combined data from two samples, in order to improve the diversity of samples, which will promisingly reflect the real clinical settings and improve the stability of the training model and the accuracy of classification. The supported vector machine method was used to construct the classification model based on LASSO features. Ten-folds CV was used to assess the reliability of the classification model. Briefly, 148 subjects were randomly separated into 10 groups. Each time, one group in turn was used as a test group and the other nine groups were used as training groups. This step was repeated 10 times, and each time, accuracy, sensitivity, specificity and recall indices were calculated. Finally, the mean of each index across the 10 times was used to assess the performance of the constructed model. The weights of each LASSO feature were also calculated to measure the importance of each feature in the classification model.

## Results

### Clinical Characteristics at Baseline

Table 1 shows the full description of demographic and clinical characteristics of patients. No significant difference was found in gender and education between responders and non-responders.

### Features after Dimensionality Reduction

Twelve features were remained in the model for combination of structural and functional MRI, involved 3 cortical features and 9 functional connections (supplementary table 2 and figure 2).

**Fig. 1.**
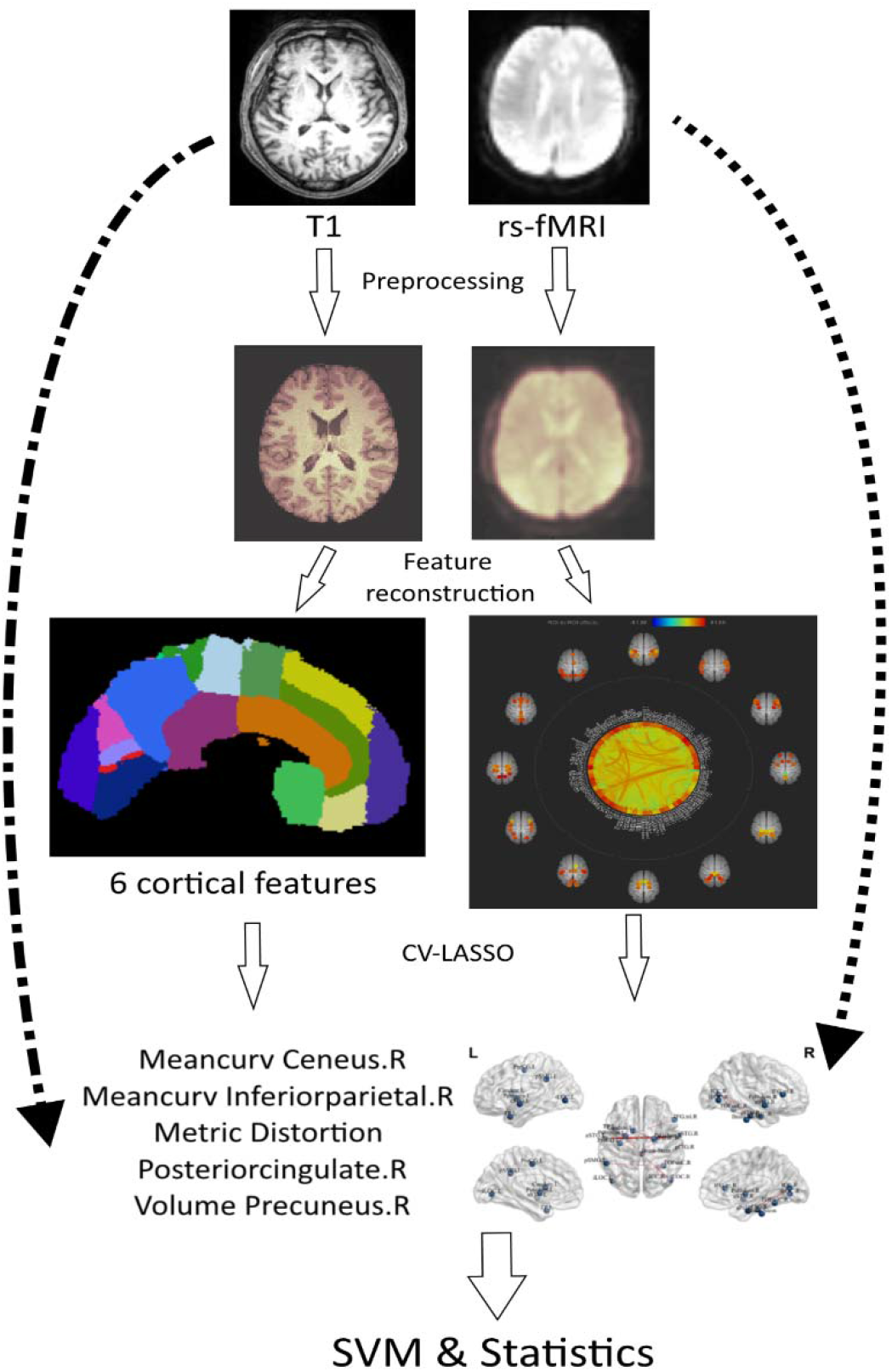
Workflow for analysis. Overview of data preprocessing, anatomical and functional feature reconstruction, feature selection, and classification.

**Fig. 2.**
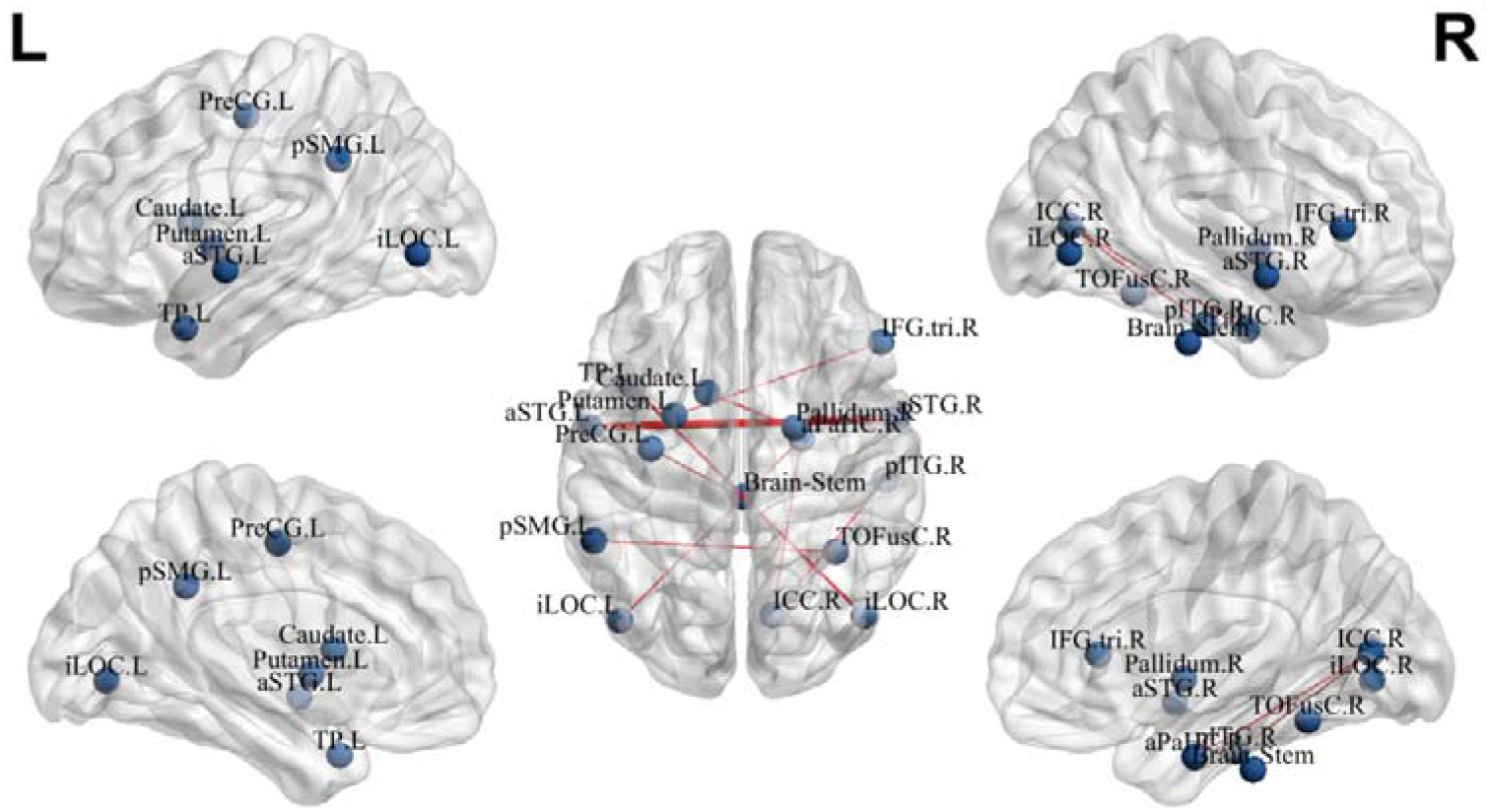
Connections contributed to prediction. Connections are scaled by their contribution degree.

### Prediction Performance

Our model combined both structural and functional features accurately predicted 92.04% responder and 80.23% non-responders to overall treatment, with an accuracy of 85.03%. The prediction accuracy was 80.38% (sensitivity: 87.28%; specificity 82.47%) for the model using functional features, and 69.68% (sensitivity: 83.96%; specificity: 72.41%) for the model using structural features (figure 3A). Results of inter-dataset cross validations confirm the capacity of discriminating responders from non-responders by features from different scanners (supplementary table 2). The analysis after excluding patients treated with ECT (18 in dataset 1; 21 in dataset 2) suggested similar predictive capacity (figure 3B). Likewise, taken rTMS into consideration, the further analysis replicated these results after excluding patients treated with rTMS (supplementary table 3). Supplementary table 4 shows the prediction performance using baseline PANSS general psychopathology score and age.

**Fig. 3.**
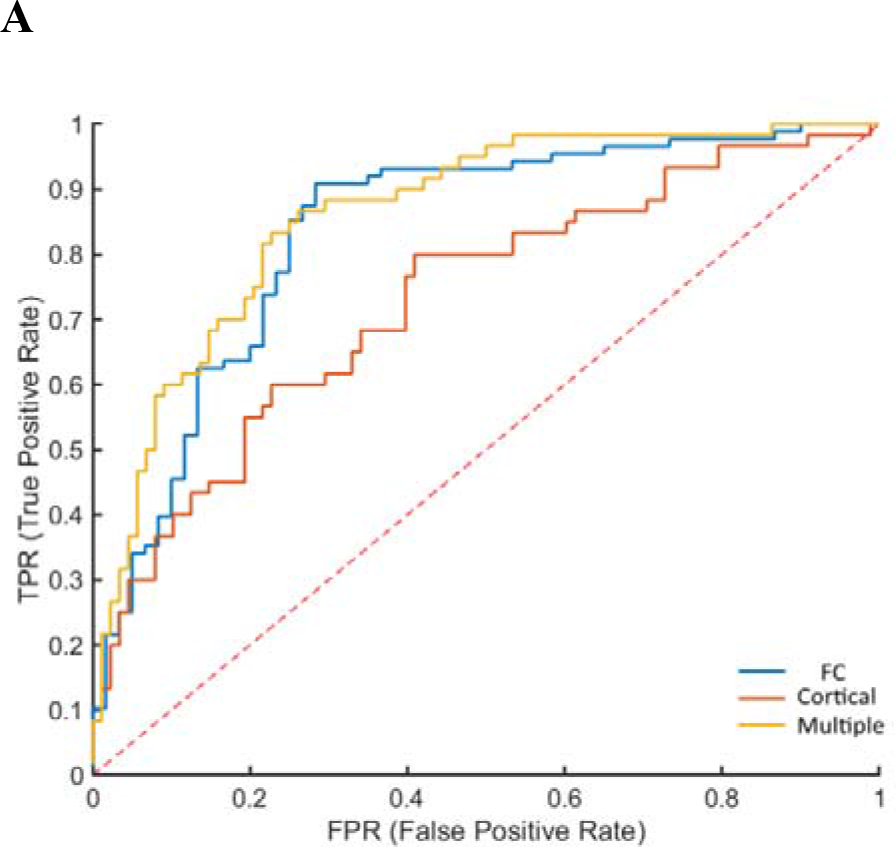
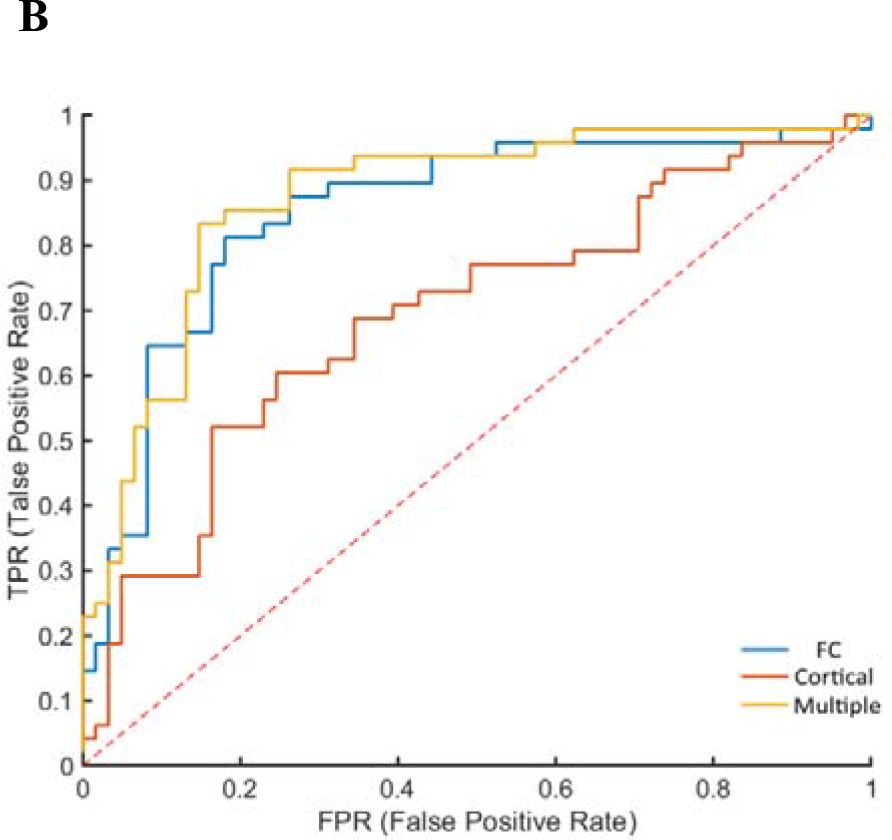
ROC curves for the prediction of treatment response with radiomics. (A) Red, blue, and yellow lines indicate predictive capacity using cortical features, functional connectivity features, and combination of them, respectively. Accordingly, areas under ROC curves were 72.97%, 83.67%, and 87.29%. (B) Red, blue, and yellow lines indicate predictive capacity using cortical features, functional connectivity features, and combination of them after excluding patients treated with ECT, respectively. Accordingly, areas under ROC curves were 69.19%, 85.14%, and 87.26%

### Correlation Analysis

No significant correlation between features and PANSS total scores survived after correction for multiple comparisons (supplementary table 5).

## Discussion

Identifying the biomarkers of schizophrenia has a critical role in fundamental research and clinical practice. In this study, combining structural/functional MRI and radiomics analysis, we effectively predict early antipsychotic treatment response in schizophrenia for the first or a single hospitalization with an accuracy of 85.03% (sensitivity: 92.04%; specificity: 80.23%), a comparable or superior capacity of MRI to that for predicting response to rTMS 5 sessions per week during the 3-week period^19^ and treatment with risperidone/aripiprazole for 12 weeks,^20^ risperidone for 10 weeks,^21^ or quetiapine for 7 months^22^ in patients with schizophrenia.

Previous studies have implicated predictive value of structural and functional MRI in schizophrenia using machine learning (supplementary table 6). Structural MRI-based markers are known to accurately predict response to rTMS in patients with schizophrenia (84.8% accuracy, 79.2% sensitivity, and 90.2% specificity), involving reduced gray matter density in the prefrontal, insular, medial temporal, and cerebellar cortices and increments in the parietal lobe and thalamus.^19^ Functional connectivity of the striatum is suggested to be a reasonable predictor for antipsychotic response (80% sensitivity and 75% specificity), showing higher response rate to be associated with lower connectivity linked to the anterior cingulate and medial prefrontal cortex and with higher connectivity linked to the posterior cerebral areas.^20^ Besides, functional connectivity of the superior temporal cortex has been found to be informative in predicting response to antipsychotics (82.5% accuracy, 88.0% sensitivity, and 76.9% specificity) using machine learning algorithms,^21^ by which the classification accuracy was 79% (sensitivity = 75%; specificity = 83%) to predict the negative symptom improvement using workig memory-related functional connectivity.^22^ While it remains to be determined which core features on structural and functional MRI possesses such a predictive capacity, an alternative approach could be the radiomics. It helps to save raw data and discover their contribution, which could be loss of certain conventional analysis.

In this study, feature selection identified cortical measures in the right precuneus, cuneus, and inferior parietal lobule and functional connectivity involved superior temporal gyrus, temporal pole, LTG, supramarginal gyrus, temporal occipital fusiform cortex, lateral occipital cortex, inferior frontal gyrus, precentral gyrus, putamen, caudate, pallidum, intracalcarine cortex, parahippocampal gyrus, and brain stem that significantly contributed to discrimination. We showed that the curvature of right inferior parietal cortex and functional connectivity between the triangular part of right inferior frontal gyrus pars and left putamen contributed mostly in the prediction, which are overlapping findings detected by above mentioned studies, and are underlying schizophrenia as demonstrated by previous studies.^23, 24^ The neuroimaging literatures have been encouraging in delving into the neural substrates behind schizophrenia.^25-28^ The current study is paving the way to establish predictive indicators based on deeper understanding of the pathophysiology of the disorder, as proposed by Tandon el al.^29^

The aim to explore neuroimaging markers of schizophrenia has long been pursued. Radiomics,^30, 31^ a central technique aiding decision making in clinical practice, is considered as “the bridge between medical imaging and personalised medicine”.^32^ A large body of studies suggests its increasing importance in medical imaging,^8^ which tries to build a deep learning algorithm for automated medicine,^33-35^ creating an ideal situation for application of deep learning in psychoradiology^36^ where there are features, rather than lesions, i.e., the alterations of cerebral structures and functions. Generally speaking, radiomics features build upon the volume of interest and then they are linked to clinical data.^37, 38^ Although no entire lesion can be used for extracting features in mental disorders, we could consider abnormal structures and functions in the brain and even the whole brain as the volume of interest for analysis, as Sun et al ^10^ and Cui et al ^9^ performed in attention deficit hyperactivity disorder and schizophrenia, respectively. These studies extracted features from the whole-brain gray matter and white matter and functional connectivity with significant difference. Holding the properties of large-scale data, both clinical imaging archives and functional imaging are reliably leveraged by radiomics method in psychiatry.

Additionally, patients suffer from numerous adverse effects induced by antipsychotics. The mean length of stay in hospital was 2-3 weeks for patients included in our study. Prediction of the very early response for the first or a single hospitalization could be helpful in implying long-term outcome of patients with schizophrenia. Radiomics methods, as demonstrated by a series of recent neuroimaging studies, hold great promise for improving the diagnosis, treatment, and prediction of prognosis in psychiatric domains, which will have an effect on personalized medicine. On the basis of neuroimaging-based markers, radiologists could provide evidence for pyschiatrists to facilitate the clinical decision making.

We need to take several considerations into account when interpreting our results. First, radiomics analysis using a large sample may increase the accuracy of predicting treatment response, but the sample size was limited after excluding patients who received ECT. Larger sample size will be helpful. Second, in addition to ECT, these two patient cohorts underwent heterogeneous treatments, which therefore may introduce an inevitable effect on the findings. Third, there is heterogeneity between dataset 1 and 2. The dataset 1 included the patients with schizophrenia, while the dataset 2 included patients with schizophrenia, schizophreniform disorder, or brief psychotic disorder. The duration varies in different disorders, and this population of patients suffered from the heterogeneous baseline severity of symptoms in dataset 1, and the patients differed regarding age in dataset 2, which might be potential confounding factors. Nevertheless, this is a very exploratory study with no hold out sample to example generalization. These concerns suggest the need for randomized controlled trials for the direct validation of radiomics in the setting of treatment response in schizophrenia.

In sum, these results highlight the power of structural and functional MRI-derived radiomics features to predict early response to treatment in schizophrenia. More expansively, our findings emphasize the predictive capacity for long-term response in schizophrenia and the clinical relevance of structural and functional neuroimaging in mental illnesses by means of radiomics, an essential strategy for individualized intervention. This work points the way toward future predictive biomarker development in the field of psychoradiology,^36^ centered on large-scale clinical imaging data, for efficient treatments reworked for now.

## Data Availability

The datasets generated for this study are available on request to the corresponding author.

## Acknowledgements

The authors acknowledge their patients and patients’ family. For advice, we thank Miss Xiao Chang (from the Department of Psychiatry, Brain Center Rudolf Magnus, University Medical Center Utrecht) and Prof. Yong He (State Key Laboratory of Cognitive Neuroscience and Learning, Beijing Normal University). The authors declare that no conflict exists.

## References

1. Fox MD. Mapping Symptoms to Brain Networks with the Human Connectome. The New England journal of medicine Dec 6 2018;379(23):2237–2245.

2. Doucet GE, Moser DA, Luber MJ, Leibu E, Frangou S. Baseline brain structural and functional predictors of clinical outcome in the early course of schizophrenia. Mol Psychiatry Oct 3 2018.

3. Cui LB, Cai M, Wang XR, et al. Prediction of early response to overall treatment for schizophrenia: A functional magnetic resonance imaging study. Brain Behav Feb 2019;9(2):e01211.

4. Rubio JM, Correll CU. Duration and Relevance of Untreated Psychiatric Disorders, 1: Psychotic Disorders. J Clin Psychiatry Mar 2017;78(3):358–359.

5. Lieberman JA, First MB. Psychotic Disorders. N Engl J Med Jul 19 2018;379(3):270–280.

6. Correll CU, Galling B, Pawar A, et al. Comparison of Early Intervention Services vs Treatment as Usual for Early-Phase Psychosis: A Systematic Review, Meta-analysis, and Meta-regression. JAMA Psychiatry Jun 1 2018;75(6):555–565.

7. Albert N, Madsen T, Nordentoft M. Early Intervention Service for Young People With Psychosis: Saving Young Lives. JAMA Psychiatry May 1 2018;75(5):427–428.

8. Gillies RJ, Kinahan PE, Hricak H. Radiomics: Images Are More than Pictures, They Are Data. Radiology Feb 2016;278(2):563–577.

9. Cui LB, Liu L, Wang HN, et al. Disease Definition for Schizophrenia by Functional Connectivity Using Radiomics Strategy. Schizophr Bull Aug 20 2018;44(5):1053–1059.

10. Sun H, Chen Y, Huang Q, et al. Psychoradiologic Utility of MR Imaging for Diagnosis of Attention Deficit Hyperactivity Disorder: A Radiomics Analysis. Radiology May 2018;287(2):620–630.

11. Jiang Y, Luo C, Li X, et al. Progressive Reduction in Gray Matter in Patients with Schizophrenia Assessed with MR Imaging by Using Causal Network Analysis. Radiology May 2018;287(2):633–642.

12. Cui LB, Wei Y, Xi YB, Griffa A, De Lange SC, Kahn RS, Yin H, Van den Heuvel MP. Connectome-Based Patterns of First-Episode Medication-Naïve Patients With Schizophrenia. Schizophr Bull Mar 30 2019.

13. Tandon R. Antipsychotics in the treatment of schizophrenia: an overview. J Clin Psychiatry 2011;72 Suppl 1:4–8.

14. Samara MT, Leucht C, Leeflang MM, et al. Early Improvement As a Predictor of Later Response to Antipsychotics in Schizophrenia: A Diagnostic Test Review. Am J Psychiatry Jul 2015;172(7):617–629.

15. Cui LB, Wang LX, Tian P, et al. Aberrant perfusion and its connectivity within default mode network of first-episode drug-naive schizophrenia patients and their unaffected first-degree relatives. Sci Rep Nov 23 2017;7(1):16201.

16. Yu H, Yan H, Wang L, et al. Five novel loci associated with antipsychotic treatment response in patients with schizophrenia: a genome-wide association study. Lancet Psychiatry Apr 2018;5(4):327–338.

17. Obermeier M, Mayr A, Schennach-Wolff R, Seemuller F, Moller HJ, Riedel M. Should the PANSS be rescaled? Schizophr Bull May 2010;36(3):455–460.

18. Cui LB, Liu L, Guo F, et al. Disturbed Brain Activity in Resting-State Networks of Patients with First-Episode Schizophrenia with Auditory Verbal Hallucinations: A Cross-sectional Functional MR Imaging Study. Radiology Jan 03 2017;283(3):810–819.

19. Koutsouleris N, Wobrock T, Guse B, et al. Predicting Response to Repetitive Transcranial Magnetic Stimulation in Patients With Schizophrenia Using Structural Magnetic Resonance Imaging: A Multisite Machine Learning Analysis. Schizophr Bull Aug 20 2018;44(5):1021–1034.

20. Sarpal DK, Argyelan M, Robinson DG, et al. Baseline Striatal Functional Connectivity as a Predictor of Response to Antipsychotic Drug Treatment. Am J Psychiatry Jan 2016;173(1):69–77.

21. Cao B, Cho RY, Chen D, Xiu M, Wang L, Soares JC, Zhang XY. Treatment response prediction and individualized identification of first-episode drug-naive schizophrenia using brain functional connectivity. Mol Psychiatry Jun 19 2018.

22. Nejad AB, Madsen KH, Ebdrup BH, Siebner HR, Rasmussen H, Aggernaes B, Glenthoj BY, Baare WF. Neural markers of negative symptom outcomes in distributed working memory brain activity of antipsychotic-naive schizophrenia patients. Int J Neuropsychopharmacol Jul 2013;16(6):1195–1204.

23. Cui LB, Liu K, Li C, et al. Putamen-related regional and network functional deficits in first-episode schizophrenia with auditory verbal hallucinations. Schizophr Res May 2016;173(1-2):13–22.

24. Huang P, Xi Y, Lu ZL, et al. Decreased bilateral thalamic gray matter volume in first-episode schizophrenia with prominent hallucinatory symptoms: A volumetric MRI study. Sci Rep 2015;5:14505.

25. Brugger SP, Howes OD. Heterogeneity and Homogeneity of Regional Brain Structure in Schizophrenia: A Meta-analysis. JAMA Psychiatry Nov 1 2017;74(11):1104–1111.

26. Palaniyappan L. Progressive cortical reorganisation: A framework for investigating structural changes in schizophrenia. Neurosci Biobehav Rev Aug 2017;79:1–13.

27. Dong D, Wang Y, Chang X, Luo C, Yao D. Dysfunction of Large-Scale Brain Networks in Schizophrenia: A Meta-analysis of Resting-State Functional Connectivity. Schizophr Bull Jan 13 2018;44(1):168–181.

28. Collin G, Keshavan MS. Connectome development and a novel extension to the neurodevelopmental model of schizophrenia. Dialogues Clin Neurosci Jun 2018;20(2):101–111.

29. Tandon N, Tandon R. Will Machine Learning Enable Us to Finally Cut the Gordian Knot of Schizophrenia. Schizophr Bull Aug 20 2018;44(5):939–941.

30. Kumar V, Gu Y, Basu S, et al. Radiomics: the process and the challenges. Magn Reson Imaging Nov 2012;30(9):1234–1248.

31. Lambin P, Rios-Velazquez E, Leijenaar R, et al. Radiomics: extracting more information from medical images using advanced feature analysis. Eur J Cancer Mar 2012;48(4):441–446.

32. Lambin P, Leijenaar RTH, Deist TM, et al. Radiomics: the bridge between medical imaging and personalized medicine. Nat Rev Clin Oncol Dec 2017;14(12):749–762.

33. Gulshan V, Peng L, Coram M, et al. Development and Validation of a Deep Learning Algorithm for Detection of Diabetic Retinopathy in Retinal Fundus Photographs. JAMA Dec 13 2016;316(22):2402–2410.

34. Esteva A, Kuprel B, Novoa RA, Ko J, Swetter SM, Blau HM, Thrun S. Dermatologist-level classification of skin cancer with deep neural networks. Nature Feb 2 2017;542(7639):115–118.

35. Hazlett HC, Gu H, Munsell BC, et al. Early brain development in infants at high risk for autism spectrum disorder. Nature Feb 15 2017;542(7641):348–351.

36. Lui S, Zhou XJ, Sweeney JA, Gong Q. Psychoradiology: The Frontier of Neuroimaging in Psychiatry. Radiology Nov 2016;281(2):357–372.

37. Aerts HJ, Velazquez ER, Leijenaar RT, et al. Decoding tumour phenotype by noninvasive imaging using a quantitative radiomics approach. Nat Commun Jun 03 2014;5:4006.

38. Adduru VR, Michael AM, Helguera M, Baum SA, Moore GJ. Leveraging Clinical Imaging Archives for Radiomics: Reliability of Automated Methods for Brain Volume Measurement. Radiology Sep 2017;284(3):862–869.

